# ExPheWas: a browser for gene-based pheWAS associations

**DOI:** 10.1101/2021.03.17.21253824

**Authors:** Marc-André Legault, Louis-Philippe Lemieux Perreault, Marie-Pierre Dubé

## Abstract

**Motivation:** The relationship between protein coding genes and phenotypes has the potential to inform on the underlying molecular function in disease etiology. We conducted a phenome-wide association study (pheWAS) of protein coding genes using a principal components analysis-based approach in the UK Biobank.

**Results:** We tested the association between 19,114 protein coding gene regions and 1,210 phenotypes including anthropometric measurements, laboratory biomarkers, cancer registry data, hospitalization and death record codes and algorithmically-defined cardiovascular outcomes. We report the pheWAS results in a user-friendly web-based browser. Taking atrial fibrillation, a common cardiac arrhythmia, as an example, ExPheWas identified genes that are known drug targets for the treatment of arrhythmias and genes involved in biological processes implicated in cardiac muscle function. We also identified *MYOT* as a possible atrial fibrillation gene.

**Availability and implementation:** The ExPheWas browser and API are available at http://exphewas.statgen.org/

## Introduction

Uncovering the phenotypic roles of human protein coding genes is an important goal of genetics to help improve our understanding of molecular physiology and disease pathology. Genetic variation provides a tool for predicting the consequences of altering the functions of a protein and is an important first step in validating drug targets for the development of therapies. Rare loss-of-function variants can be used to assess the impact of disrupting protein function, but they are frequently untyped in large population cohorts that rely on genotyping platforms. In such cohorts, it may be possible to use many common variants with small effects to establish the association between a locus and phenotype. Gene-based pheWAS analysis can also support annotation efforts as follow-up to a GWAS discovery (Denny *et al*., 2010).

Many computational approaches to collapse common variants into gene-based association tests have been developed and an overview of methods is shown in Table 1. In general, one of the major challenges is to account for the correlation between genetic variants due to linkage disequilibrium (LD) that induces correlation between the marginal association statistics. Methods have heterogeneous performance with respect to the underlying genetic architecture, the inclusion of common and rare variants and the presence of interactions (Wu *et al*., 2011; Wang *et al*., 2017). The required input may also be a concern as some methods require individual-level genotypes (*e*.*g*. SKAT or PLINK SNP set) whereas others rely on summary association statistics and a population-matched LD reference panel (*e*.*g*. VEGAS or GATES). Computational burden also varies with algorithms relying on permutation tests to derive empirical p-value typically being slower (Li *et al*., 2011). Among popular techniques are the use of a simulation-based approach (Liu *et al*., 2010), kernel-based association tests (Ionita-Laza *et al*., 2013) and principal component analysis (PCA)-based methods (de Leeuw *et al*., 2015). More details and flexible software packages implementing different methods have been previously published (Svishcheva *et al*., 2019). Using a gene-based association testing approach reduces the multiple hypothesis testing burden and facilitates downstream bioinformatics analyses that require gene-level annotations, such as gene set enrichment analysis.

**Table 1.** Summary of gene or locus-based association tests. A brief description of the methods is provided along with major characteristics. The computational cost is given as a qualitative estimate and accounts for the computation of single variant association statistics for tests that rely on them.

| Name | Description | Allows weighting | Used with summary statistics | Genetic variant frequency | Comp. cost | Method to account for LD | Reference |
| --- | --- | --- | --- | --- | --- | --- | --- |
| PLINK Set-based test | Mean of $\chi^2$ statistics amongst strongest associations | No | No | Common | +++ | Empirical P value from phenotype permutations | (Purcell *et al.*, 2007) |
| Versatile gene-based association study (VEGAS) | Sum of $\chi^2$ statistics from single-variant models | No | Yes | Common | ++ | Simulates association statistics under the null using the LD matrix<br>Empirical P-value computation | (Liu *et al.*, 2010) |
| GATES | Adaptation of the Simes test for multiple testing correction<br>Estimates the effective number of P-values (tests) from their correlation matrix | Yes | Yes | Common | + | Estimation of the effective number of tests in the P-value correction | (Li *et al.*, 2011) |
| Burden tests | Test based on the count or distribution of alternative alleles | Yes | No* | Rare | - | None (no strong impact for rare variants) | Reviewed in (Lee *et al.*, 2014). Popular implementation described in (Zhan *et al.*, 2016) |
| Kernel-based tests (e.g. SKAT) | The kernel function can be seen as a scalar genotypic similarity measure for pairs of individuals allowing for complex relationships<br>Semi-parametric models are use for statistical testing | Yes | No* | Common and rare | + | No formal modeling in LD<br>Depends on the choice of kernel | (Kwee *et al.*, 2008; Wu *et al.*, 2011) |
| Combined Association Test (COMBAT) | Ensemble learning approach to combine results from different methods | - | Yes | Common | ++ | - | (Wang *et al.*, 2017) |
| Principal Component Analysis (PCA)-based methods | Uses PCA to construct linear combinations of genetic variants that form an orthogonal basis<br>Inference is based on testing for the gain in goodness of fit from including principal components in conventional regression models | No** | No* | Common | - | LD is accounted for because the principal components are orthogonal | (Gauderman *et al.*, 2007; de Leeuw *et al.*, 2015) |
| PASCAL | Optimized implementation of tests based on the mean and maximum $\chi^2$ statistics<br>P-value is computed using Monte Carlo simulation or from an optimized numerical integration algorithm | No | Yes | Common | + | LD is modeled through the correlation matrix of summary statistics | (Lamparter *et al.*, 2016) |

\* Even though it was not possible in their original description, the tool SUMmary statistics Fast REGional Association Tests provides implementations for these algorithms that allow for use with summary statistics (Svishcheva *et al.*, 2019).
\*\* Weighting may be possible as is done in PCA-based Mendelian randomization methods (Burgess *et al.*, 2017)

Here, we implemented an efficient PCA-based association testing method suitable for pheWAS analysis of protein coding genes in the UK Biobank. Our phenome-wide scan considered anthropomorphic measurements, laboratory measurements, hospitalization or death codes, algorithmically-defined cardiovascular outcomes as well as self-reported diseases. We developed a web-based results browser available at http://exphewas.statgen.org/ and provide programmatic access through a publicly available API. We also characterize our association testing strategy in terms of power and hyperparameter sensitivity by using well-known drug target genes as examples. Finally, we used atrial fibrillation, a common cardiac arrhythmia to demonstrate how our resource may be used in practice.

## Methods

### UK Biobank and genetic quality control

The UK Biobank is a longitudinal population cohort of more than 500,000 individuals. All participants visited a recruitment center between 2006 and 2010 where urine and blood samples were collected allowing for the measurement of an extensive panel of biochemical markers. A touchscreen-based questionnaire was also filled and followed-up with a verbal interview with a nurse allowing participants to self-report a wide variety of diseases. Genetic data derived from a genome-wide genotyping array was also collected and imputed to about 96 millions genetic variants (Bycroft *et al*., 2018).

Because of the high throughput nature of our study, we conducted a strict genetic quality control to reduce the risk of bias due to poor genotyping or imputation quality or population stratification. We excluded all variants and individuals with more than 2% of data values missing. To avoid bias due to cryptic relatedness, we also randomly selected one individual from pairs predicted to be related using a kinship coefficient corresponding to a 3^rd^ degree relationship (0.0884) as a threshold. Our analysis also included only individuals from the largest genetically homogeneous population in the UK Biobank corresponding to individuals of European ancestry. We excluded individuals from different ancestry based on self-reported data or outliers from a manually defined cluster in the genome-wide PCA plot. After these steps, a total of 413,138 individuals remained for analysis.

### Creation of gene-based PCs

To create a compact representation capturing genetic variability within autosomal gene regions, we conducted PCA with genetic variants of minor allele frequency (MAF) of 0.01 or greater at every locus. We used the Ensembl gene boundaries and added a padding of 3 kilobases (gene boundaries ± 1.5 kilobases), extracted all additively encoded genotype dosages and conducted the PCA using the implementation from *scikit-learn* in the Python programming language (Pedregosa *et al*., 2011). The projection on the space spanned by the PCs was saved for all samples.

We excluded genes located on sexual chromosomes because systematic encoding differences between men and women risked creating spurious associations. For example, using the gene *ACE2* located on the X chromosome, we used a logistic regression model to test if the first 11 PCs, explaining 95% of the variance in the genotypes, could predict the individual’s sex. The log-likelihood ratio test of the joint effect of the 11 included PCs, had a p-value of 0.032 suggesting that the gene-based PCs were different between men and women. The sex-stratified computation of gene-based PCs is likely to address this problem and will be considered for future releases.

### Association testing

Different approaches have been developed for association testing of common variants at a gene locus (Svishcheva *et al*., 2019; de Leeuw *et al*., 2015; Liu *et al*., 2010). Because our application, a pheWAS, required a large number of tests, we focused on an approach using dimensionality reduction as a first step to reduce the computational burden. We opted for a principal component analysis-based association model that was proposed in the MAGMA software (de Leeuw *et al*., 2015; Gauderman *et al*., 2007). We also propose an extension of the method based on the analysis of deviance which can be used for non-continuous outcomes.

This approach is valid for any model using maximum likelihood estimation including generalized linear models.

For continuous traits, an F test is used to compare two nested models as in MAGMA. First, a null model regresses the outcome on covariates such as age, sex and genome-wide principal components to adjust for confounding due to ethnicity. The association test is based on the gain in goodness of fit (or lack thereof) after adding gene-based PCs to the null model. More formally, given two nested models with *P*_*1*_ and *P*_*2*_ representing the set of parameters where *P*_1_ ⊂ *P*_2_, then one can compute the F statistic given by

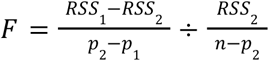

Where RSS_i_ and p_i_ are the residual sum of squares and number of parameters of the i^th^ model, respectively and *n* is the number of samples. This statistic follows an *F* distribution with (p_2_-p_1_, n-p_2_) degrees of freedom under the null hypothesis that the second model does not improve the residual sum of squares.

A similar approach for generalized linear models has been described based on the difference in deviance and was termed the analysis of deviance (McCullagh and Nelder, 1983). The difference between the deviances of the nested models follows a χ^2^ distribution with p_2_-p_1_ degrees of freedom under the null hypothesis. This extension can be used in the context of logistic regression as used in our analyses. In this setting, an equivalent approach based on the likelihood ratio test was also previously suggested (Gauderman *et al*., 2007).

Both the F-test and analysis of deviance based methods were implemented in R under the “anova” function that takes fitted models as parameters (*i*.*e*. output from the *lm* or *glm* functions). Because of the large number of tests in our analysis, we used the optimized *fastglm* (v0.0.1) package when fitting logistic regression models for binary outcomes (https://github.com/jaredhuling/fastglm). This package uses the Cholesky decomposition to optimize the iterative reweighted least squares algorithm which drastically reduced the time required for logistic regressions.

In the pheWAS study, we repeated the association tests about 25 million times. This certainly results in false positives and we used a false discovery rate (FDR) approach to control for multiple hypothesis testing. As we believe that considering all tests together is not suitable for most research questions, we integrated a FDR mechanism on a per gene or per phenotype basis. Specifically, we used the test p-values to compute corresponding *q* values designed so that if all *q* values ≤ 0.05 are considered significant, then 5% of the significant features will be false discoveries on average (Storey and Tibshirani, 2003). For example, when browsing results for a gene of interest, the displayed *q*-values will be based on the 1,127 tested phenotypes. Browsing the same results by “outcome” will result in a different *q*-values because the latter would be based on all tested genes in association with the outcome of interest. In all cases, the uncorrected p-value and the Bonferroni corrected p-value are also provided. When browsing associations for a given gene, a quantile-quantile plot of association p-values is also displayed on the results browser along with the λ inflation factor corresponding to the ratio of the median observed association statistic to the median expected association statistic. The inflation factor is a quantitative measurement of deviation from the null and high λ (above one) values represent loci that contribute to many phenotypes (*i*.*e*. pleiotropic loci). For example, the *HLA-DQB1* gene is notoriously pleiotropic and has λ=2.48.

In a sensitivity analysis exploring the optimal threshold for the variance explained for selected drug target genes, we repeated the association test while varying the number of included PCs. When the association p-values were numerically close to zero for many choices of PCs, we randomly selected a subset of individuals to improve the resolution.

### Power analyses

We simulated continuous phenotypes for every simulation replicate using two models representing a scenario with a single causal variant and a scenario where all variants contribute to the heritability in a random effects model. For the single causal variant model, we randomly sampled a causal variant and fixed its effect coefficient (*β*) as a simulation parameter. We then added gaussian noise so that the total variance in the continuous simulated phenotype was 1. In the second model, every variant had an effect sampled from a standard normal distribution and gaussian noise was added to achieve the heritability (h^2^) set as a simulation parameter (Yang *et al*., 2010). The simulations used real genotype data from the *PCSK9* locus in the UK Biobank containing 138 common variants as a typical example of phenotype-associated protein coding gene of median length. We randomly selected 20,000 individuals and repeated the simulation procedure 1,000 times. For simulated phenotypes, we tested the association between every genotype at the locus and the simulated phenotype using linear regression. We estimated the power for the conventional linear regression model as the fraction of simulation replicates where the minimum variant p-value was under a bonferroni corrected threshold of 0.05 / 138. For the gene-based association test, we used the fraction of F tests with a p-value under 0.05 across simulation replicates.

### PheWAS analysis

For every gene, we tested the joint association of principal components explaining 95% of the genetic variance at the gene locus with 83 continuous phenotypes and 1,127 binary phenotypes. The continuous phenotypes include anthropometric measurements (such as height or body mass index), laboratory measurements (such as lipoprotein levels, glycemic traits or blood cell traits) and imaging measurements (such as the intima-media thickness). Due to the high throughput approach used, we manually transformed continuous variables so that they were approximately normally distributed. This step was taken to avoid violating the assumption of normally distributed residuals in linear regression as it was not feasible to run conventional model diagnostics for all gene-phenotype pairs individually. The description of the included continuous variables along with the transformation used, if needed, is presented in Supplementary Table 1.

For the binary phenotypes, we used the self-reported diseases from the UK Biobank and the ICD10 codes reported in the hospitalization or death records. We also grouped the ICD10 codes according to two hierarchical levels to represent increasingly broad disease definitions. Specifically, we tested ICD10 blocks (*e*.*g*. I10-I15 Hypertensive diseases) as well as more precise 3 character codes (*e*.*g*. I11 Hypertensive heart disease). For cancer data, the UK Biobank also provides linkage with a national cancer registry. For all ICD10 codes corresponding to neoplasms (C00-D49), we used the cancer registry data instead of the hospitalization data. Because of the shared etiology between cancer subtypes, we also excluded individuals with any cancer from being controls for analyses of other cancer subtypes. For hospitalization or death records, the initial analysis (data release v0.1) included hospitalization records up to 2016-03-13 and the latest recorded date of death was 2016-02-16. Codes from either the primary or secondary codes were used in the pheWAS to maximize the number of cases and statistical power.

We also included algorithmically-defined cardiovascular outcomes as phenotypes in the pheWAS. This includes coronary artery disease, angina, heart failure, myocardial infarction and coronary revascularization procedures (percutaneous coronary intervention and coronary artery bypass graft). The codes and procedure used to define these variables is presented in Supplementary Table 2.

To efficiently conduct the PheWAS analysis of the previously described gene-based PCs and phenotypes, we used a custom R script (tested under R v3.6.0) that is available online (https://github.com/legaultmarc/UKBPheWAS).

### Interface and API

To present the results of the PheWAS, we developed an API and a web-based results browser. The Python programming language (v3.8) and Flask web framework (v1.1.1) were used to develop both. We used the SQLAlchemy (v1.3.13) object relational mapper and SQL engine to build the results database and the publicly available instance uses PostgreSQL (v9.6.3). The code for the database models, API endpoints, web browser endpoints and the JavaScript frontend are publicly available online https://github.com/pgxcentre/ExPheWAS. Interactive visualizations integrated in the web application were developed using D3.js (https://d3js.org/). These include median tissue expression of genes in GTEx V8 used to contextualize the expression patterns of the tested protein coding loci as well as the drug target enrichment analyses.

To ensure long term availability, the service is hosted on the Compute Canada Cloud (https://www.computecanada.ca/home/).

### Enrichment analyses

We integrated enrichment analysis utilities to the API and results browser. For a given phenotype, it is possible to test if the associated genes are enriched in drug targets. To achieve this, we used the ChEMBL database to map drug target genes to Anatomical Therapeutic Chemical Classification (ATC) codes representing drug classes (Bento *et al*., 2014). The ATC codes are structured hierarchically in a 5 level system where the first level indicates the anatomical group (*e*.*g*. C represents drugs acting on the “cardiovascular system”) and the fifth level represents individual drugs (*e*.*g*. C07AB07 represents bisoprolol, a specific beta-blocker molecule). For enrichment analyses, we used two complementary approaches. The first approach is based on a Fisher exact test of the 2×2 contingency table of the number of genes associated with the phenotype and drug class at a q-value ≤ 0.05 level. We also provided results from a *Fast Gene Set Enrichment Analysis* (FGSEA) implementation where ATC codes are treated as pathways and the association statistics are ranked to evaluate enrichment (Korotkevich *et al*., 2019; Subramanian *et al*., 2005). One advantage of this approach over the Fisher exact test is that it does not categorize genes as associated or not based on a *q*-value threshold.

The results of enrichment analyses are displayed on the ExPheWas browser results page as an interactive tree with collapsible nodes. A color scale is used to represent enrichment p-value. For every ATC node, the fill color represents the node’s enrichment p-value and the stroke color represents the minimum p-value in the subtree rooted at the current node.

For the atrial fibrillation example, we also tested enrichment of various ontology terms within the genes associated with atrial fibrillation with q ≤ 0.01 (137 genes). The tested ontologies included the Gene Ontology, the KEGG pathways, and the Human Phenotype Ontology. This analysis was done using the g:Profiler web-based tool (Raudvere *et al*., 2019).

## Results

### Gene PCA

We used the Ensembl 87 database as a reference for the human protein coding genes. Out of the 20,356 genes, 926 were not on autosomes and another 316 genes either had no common variants in the UK biobank imputed dataset or did not converge because of unreasonable memory requirements during association testing. In total, 19,114 genes on autosomal chromosomes were included in the final analysis.

To efficiently represent the majority of the genetic variation at every protein coding locus, we used PCA of the additively encoded genotypes and retained the number of principal components necessary to explain 95% of the total variance. As a sensitivity analysis, and for a subset of the genes, we also tested the inclusion of PCs explaining 99% of the total variance. Results for both analyses are presented in the ExPheWas browser. The first principal components generally assigned more weight to genetic variants that are highly correlated to many other variants (*i*.*e*. good “tagging” variants). Supplementary Figure 1 shows a representative example of this pattern. We also note that the principal components (PCs) explaining a smaller portion of the variance (later components) tended to attribute larger weights to individual variants that are less correlated with other variants (smaller LD score). The number of principal components required to capture 95% of the genetic variance at a gene region varied greatly between genes, and was largely determined by the size of the region and LD structure.

### Validation of the association testing approach

#### Marginal association of PCs

To gain insight into the effect of PCs on phenotypes, we used the well known example of the proprotein convertase subtilisin/kexin type 9 (PCSK9), a protein implicated in the recycling of low density lipoprotein (LDL) receptor and consequently of LDL cholesterol levels and coronary artery disease (CAD) (Cohen *et al*., 2006). When regressing individual *PCSK9* PCs in univariate models with LDL cholesterol and CAD, we observe that the first PCs have lower standard errors as they capture the overall effect of many common variants with small weights (Supplementary Figure 2). Subsequent PCs that explain a smaller proportion of the variance and rely more on individual variants in low LD have larger standard errors, but may still have strong effects on the phenotypes. However, these effects may rely on individual variants for which traditional association tests may be more powerful. For example, PC14 in PCSK9 is strongly associated with LDL-c levels (*β* = 0.038, 95% CI [0.035, 0.041] mmol/l, p = 2 × 10^−147^) and CAD (*β* = 0.026, 95% CI [0.015, 0.037] in log odds, p = 2 × 10^−6^) despite explaining 0.86% of the overall variance in genotypes. For this PC, the variant rs11591147 is an outlier of the component weight distribution and this missense *PCSK9* variant has a strong effect on LDL-c in the GLGC (*β* = 0.50 [0.46, 0.53], p = 9 × 10^−143^) and coronary artery disease (allelic *β* = 0.50, 95% CI [0.46, 0.53], p = 9 × 10^−143^) in the CARDIoGRAMplusC4D consortium (allelic OR 1.29, 95% CI [1.16, 1.45], p = 7 × 10^−6^) (Global Lipids Genetics Consortium *et al*., 2013; Nikpay *et al*., 2015).

It is also noticeable that some PCs are positively correlated with partial gain of function whereas others represent partial loss of function as can be seen by the positive and negative effects in Supplementary Figure 2. Finally, in the example of PCSK9 the first PCs have small effect sizes on LDL cholesterol and CAD. This pattern is not systematically observed with other genes as it may depend on LD structure and selective pressures. For example, we observed the opposite trend when considering the *CETP* gene encoding the cholesteryl ester transfer protein (CETP) a well known drug target associated with high density lipoprotein cholesterol levels. For this gene, the first PCs have among the strongest effects of all individual components (Supplementary Figure 3).

#### Joint association testing of PC

The association model we used requires the selection of the number of PCs to include. This choice is important as including PCs with no phenotypic effect will result in decreased statistical power. If there is a single gene of interest, it is possible to tune this hyperparameter using cross-validation, but for a pheWAS approach and when the associated phenotype is not known a priori, using a fixed threshold is necessary.

To assess power with respect to the number of included PCs, we used drug targets that are known to be associated with various continuous or discrete phenotypes (Supplementary Table 3). We assessed the association p-value based on an increasing number of included PCs in association tests with selected phenotypes (Supplementary Figure 4). There was no best choice in the number of PCs to include to maximize power that was shared for all genes. For example, a single PC explaining 36% of the variance maximizes power for the association between *HMGCR* and LDL cholesterol with decreasing benefit afterwards. On the contrary, power is maximized after including 29 PCs explaining 92% of the variance for *GLP1R* and glycated haemoglobin.

To increase our chance of detecting associations driven by less common variants and to avoid false negatives, we selected the threshold of 95% of the variance explained for the pheWAS analysis in the UK Biobank.

#### Power analyses

The first stop for many investigators in search of gene-phenotype associations are online catalogs such as the OpenTarget genetics platform, the GWAS catalog, the SAIGE-based PheWeb platform and PhenoScanner (Gagliano Taliun *et al*., 2020; Kamat *et al*., 2019; Ochoa *et al*., 2021). These portals mostly focus on reporting variant-based association statistics and so the strongest variant-phenotype associations within a gene are typically of interest. For this reason, gene-level association testing is most commonly based on using the minimum association p-value within the gene boundaries. As we aim to provide a complementary resource to these portals, we compared the behaviour of the PC-based association method to this approach in terms of power and false positive rate.

In a scenario where there is a single causal variant, the minimum p-value in the region approach is more powerful (Supplementary Figure 5). However, under a random effects model where all variants make small contributions to the heritability, the PC-based approach is slightly more powerful and the estimated power was numerically higher for the PC-based approach for 97% of the simulated h^2^ values (Supplementary Figure 5). When the simulated heritability was zero (null model of no genetic effect), the fraction of simulation replicates where the null was rejected was 3.6% (95% CI 2.4%, 3.9%) and 2.9% (95% CI 1.9%, 3.9%) for the PC-based approach and minimum linear regression p-value approach, respectively. In both cases these false positive rates were close to the nominal levels (α = 0.05). The lower false positive rate for the minimum linear regression p-value approach was likely due to the conservative Bonferroni correction which does not account for linkage disequilibrium.

#### PheWAS

To estimate the association of human protein coding genes with multiple phenotypes, we used a phenome-wide association study approach (Denny *et al*., 2010). We tested the association between 19,114 protein coding gene regions and 1,210 phenotypes in the UK Biobank for a total of about 23 million tests (Figure 1). Some of these tests however are redundant as phenotypes may be correlated and genomic regions may overlap.

**Figure 1.**
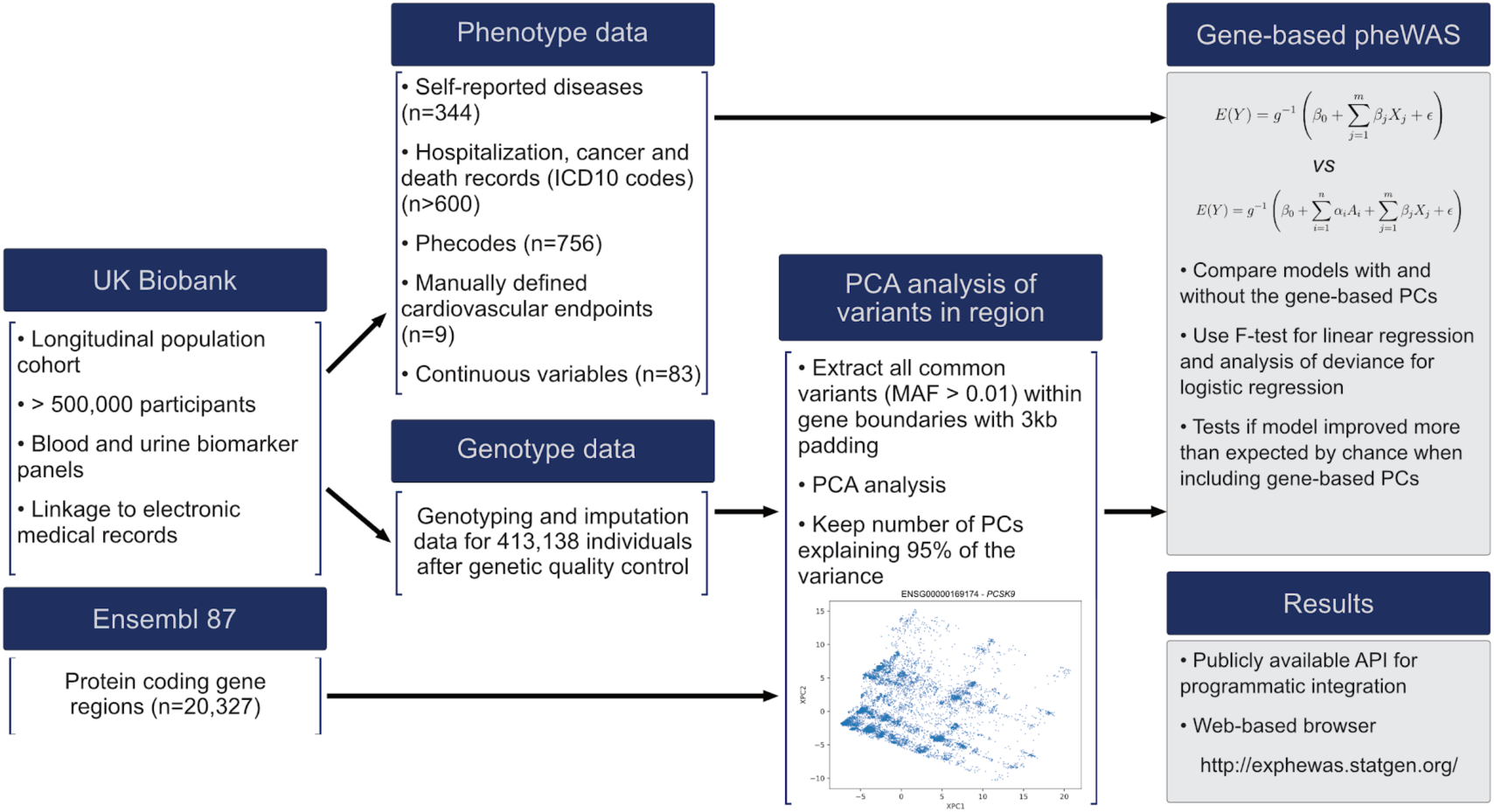
Schematic overview of the analysis from data sources to pheWAS results. Contributions to this study: the results of the gene-based pheWAS and the web-browser are represented by shaded boxes. In the equations detailing the association model, *β*_j_ terms represent m included covariables X_j_, and the α_i_ terms represent the fixed effects for n included gene-based PCs A_i_. The g^-1^ function represents the generalized linear model link function (the identity for linear regression and the logit for logistic regression) and ∈ represents the error term. The association test measures the improvement of goodness of fit when including the n gene-based PCs to the model.

We also provide information to enable the assessment of the statistical significance of associations, including the raw p-value, the conservative Bonferroni p-value and the q-value that is related to the false discovery rate. When browsing results for a gene, the q-value will account for all tested phenotypes in association with that gene. When browsing results for a phenotype, then the q-value will control for all tested genes in association with that phenotype. This enables multiple testing adjustments for the specific questions: “what genes are associated with this phenotype?” or “what phenotypes are associated with this gene?”.

#### Application programming interface and web interface

The pheWAS results consist of a very large collection of association statistics between the tested gene loci and phenotypes. These results may be useful in a wide range of applications and to help answer different research questions, making the development of a convenient results browser important. We opted to offer both an application programming interface (API) to facilitate integration with other platforms or bioinformatics resources as well as a web-based interface to allow researchers to interactively browse the results. A summary of the data available in the ExPheWas browser along with representative visualizations are presented in Figure 2. The main web-page to access these resources is http://exphewas.statgen.org. API documentation is provided along with examples.

**Figure 2.**
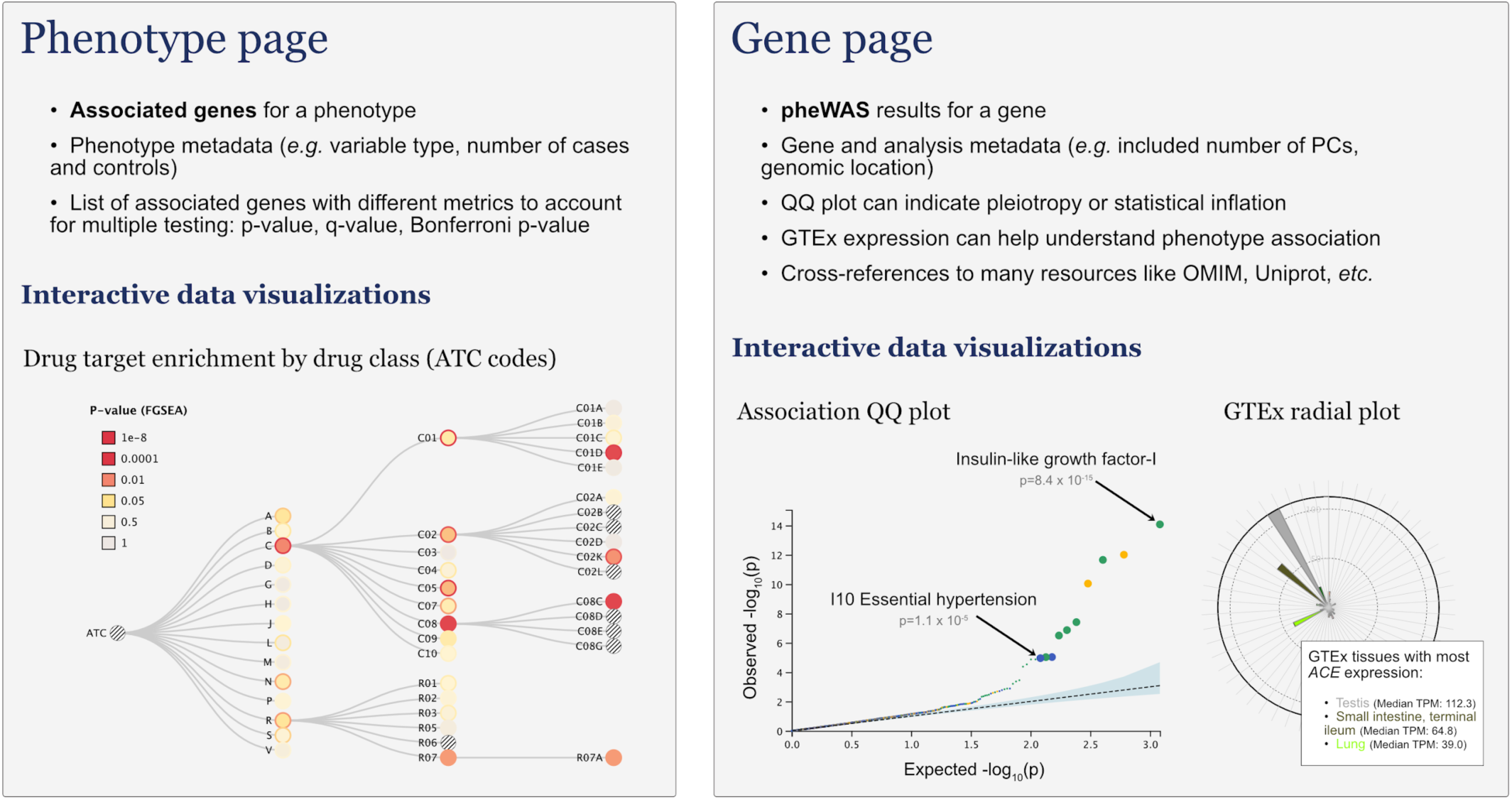
Summary of the data presented in the ExPheWas browser and adapted data visualization. Left. Results presented on the phenotype results page. The example for the gene-set enrichment analysis is for the I10 Essential (primary) hypertension code. Enriched drug classes include C01D (vasodilators used in cardiac diseases, p=0.03), C02K (other antihypertensives, p=0.016), C08C (selective calcium channel blockers with mainly vascular effects, p=0.002), R07 (other respiratory system products, p=0.019). **Right**. Results presented on the gene results page. The example data visualisations are for the angiotensin I converting enzyme (ACE gene, ENSG00000159640). The leftmost plot is a QQ plot representing deviation from the expected distribution of p-values under the null hypothesis. The statistical inflation as measured by genomic control is λ=1.13 (ratio of observed to expected median values on the QQ plot). The top 10 associated phenotypes were magnified and two were identified to emulate the interactivity available on the web version. The rightmost plot is a radial plot of median expression in GTEx v8 tissues which can be used to contextualize association findings.

## Real data application

Atrial fibrillation is a common cardiac arrhythmia of the atria which can lead to severe complications including stroke, cardiomyopathy and heart failure (Rahman *et al*., 2014; Qin *et al*., 2019). Recent GWAS including more than a million individuals have identified a large number of loci associated with atrial fibrillation (Roselli *et al*., 2018; Nielsen *et al*., 2018).

Our pheWAS study (data release v0.1) included 14,747 UK Biobank participants with evidence of atrial fibrillation in their hospitalization or death records (I48 ICD10 code, “Atrial fibrillation and flutter”). There were 137 genes associated with atrial fibrillation with a q-value ≤ 0.01. Out of these genes, 37 were also reported as the mapped gene (including upstream or downstream genes for intergenic loci) in GWAS Catalog associations (Welter *et al*., 2014). We used g:Profiler to conduct an ontological term enrichment analysis of the 137 genes and found enrichment for supraventricular arrhythmia (Human Phenotype, HP:0005115; p_adj_ = 3.7 × 10^−7^), cardiac muscle contraction (Gene Ontology Biological Process, GO:0060048; p_adj_ = 9.5 × 10^−6^) and Z disk (Gene Ontology Cellular Component, GO:0030018; p_adj_ = 0.001) among other relevant terms (Supplementary Figure 6, Supplementary Table 4).

We also tested the enrichment of atrial fibrillation associated genes in drug targets based on the ChEMBL database (Bento *et al*., 2014) as implemented in the ExPheWas browser. We found enrichment for class Ia and class Ib antiarrhythmics corresponding to ATC codes C01BA and C01BB, respectively. The Fisher’s exact test enrichment p-values for these classes were 0.006 for both. This finding is concordant with pharmacological treatment of atrial fibrillation suggesting genetic support for these drug targets. However, there was also an enrichment with local anesthetics belonging to ATC code C05AD which may represent spurious associations driven by sodium channels genes including *SCN5A* and *SCN10A* both of which are robustly associated with atrial fibrillation (McNair *et al*., 2004).

We further explored gene associations that were not previously reported in the GWAS Catalog. Notably, myotilin encoded by the *MYOT* gene was associated with heart rate (p=8.6 × 10^−31^), and atrial fibrillation (p=4.9 × 10^−11^) in our pheWAS. This region is located in a long LD block spanning multiple genes (Supplementary Figure 7, panel A.). Other credible genes that could drive the association signal in this region are *FAM13B* (atrial fibrillation p=1.9 × 10^−9^), *PKD2L2* (p=3.1 × 10^−9^), *WNT8A* (p=2.1 × 10^−11^) and *NME5* (p=5.7 × 10^−6^). The two top genes according to our analysis are *MYOT* and *WNT8A* which have p-values two orders of magnitude smaller than the others. However, *WNT8A* is not expressed in heart tissues in GTEx whereas *MYOT* is expressed in the heart. Myotilin is a component of the sarcomeric Z-disk, a structure implicated in muscle contraction and rare mutations in myotilin cause myofibrillar myopathy which often co-occurs with cardiomyopathy (OMIM:604103, (Selcen and Engel, 2004; McKusick, 1998)). In single-cell RNA sequencing analysis, *MYOT* was found to be particularly expressed in left atrial and right ventricular cardiomyocytes in line with atrial fibrillation pathophysiology (Tucker *et al*., 2020). Even though it was located nearby, we did not consider *KLHL3* as it was revealed to represent an independent association signal through stepwise conditional analysis (Supplementary Figure 7, panel B.). After two stages of the forward conditional stepwise analysis, no significant variants remained associated (Supplementary Figure 7, panel C).

## Discussion

We used a PC-based association approach to test the association between all human protein coding loci and an exhaustive set of phenotypes in a gene-based pheWAS approach. This allowed us to build a web-based resource that provides a complementary repository to current databases providing gene to phenotype mappings. Using a gene-based approach favors analyses that rely on gene-level annotations such as drug target validation or ontological enrichment analyses. There was an important computational advantage in using a PCA-based approach for our pheWAS as it greatly reduced the number of models to fit when compared to variant-based approaches. We also evaluated the power of PC-based approaches compared to conventional association testing, characterized the behaviour of individual PCs in marginal models and evaluated the effect of the choice of included PCs on the statistical association. These auxiliary results provide insight that may be useful for the development and improvement of PC-based methods in association testing and in other contexts such as Mendelian randomization (Burgess *et al*., 2017). Finally, we evaluated our gene-based pheWAS resource using a real world example by exploring genes associated with atrial fibrillation. We identified 137 atrial fibrillation-associated genes at a FDR of 0.01 and these genes were enriched for the targets of drugs used to treat atrial fibrillation and GO terms of heart development and physiology. We also focused on previously unreported genes that could be associated with atrial fibrillation and prioritized *MYOT* as an interesting candidate.

There are some limitations to our approach. First, we used gene boundaries and 3 kb of padding to define protein coding loci used for association analysis, but there is no guarantee that the association is driven by the gene product of interest and not by overlapping genes or other DNA elements. In regions of high linkage disequilibrium, it is also possible that an association is in fact due to correlation with a neighboring gene.

These limitations are shared by most association testing approaches and can only be addressed by careful interrogation of candidate associations. Power was maximized when common variants in a gene made small contributions to the phenotype as opposed to when the effect is driven by a single causal variant. This is to be expected as the strength of our approach is in the collapsing of variants at a locus. Because of the multivariable nature of the association model, there is also no intuitive measure of effect size for which conventional methods based on single variants may be more appropriate.

To conclude we contribute an important atlas of gene to phenotype associations along with tools to interrogate, contextualize and interpret the results. We believe that dimensionality reduction approaches such as PCA provide a natural way of addressing linkage disequilibrium in statistical genetics models that aggregate common variants. With the increasing number of large cohorts with available genotype data, it is essential to further develop gene-based methods that aggregate common variants with weak effects to complement other approaches such as the analysis of loss-of-function mutations.

## Data Availability

Access to the UK Biobank resource requires application through the Access Management System and instruction are available online: https://www.ukbiobank.ac.uk/enable-your-research/apply-for-access.
The ExPheWas results browser is available online at https://exphewas.statgen.org/ and instructions for programmatic access through the API are provided in the documentation section. The code for the results browser including the database models, the web application and the data visualizations is open source and available at https://github.com/pgxcentre/ExPheWAS.

https://exphewas.statgen.org/

https://github.com/legaultmarc/UKBPheWAS

## Acknowledgements

We thank the UK Biobank for providing the data under Application Number 20168.

## Data Availability

Access to the UK Biobank resource requires application through the Access Management System and instructions are available online: https://www.ukbiobank.ac.uk/enable-your-research/apply-for-access.

The ExPheWas results browser is available online at https://exphewas.statgen.org/ and instructions for programmatic access through the API are provided in the documentation section. The code for the results browser including the database models, the web application and the data visualizations is open source and available at https://github.com/pgxcentre/ExPheWAS.

## References

Bento, A.P. et al. (2014) The ChEMBL bioactivity database: an update. Nucleic Acids Res., 42, D1083–90.

Burgess, S. et al. (2017) Mendelian randomization with fine-mapped genetic data: Choosing from large numbers of correlated instrumental variables. Genet. Epidemiol., 41, 714–725.

Bycroft, C. et al. (2018) The UK Biobank resource with deep phenotyping and genomic data. Nature, 562, 203–209.

Cohen, J.C. et al. (2006) Sequence Variations in PCSK9, Low LDL, and Protection against Coronary Heart Disease. N. Engl. J. Med., 354, 1264–1272.

Denny, J.C. et al. (2010) PheWAS: demonstrating the feasibility of a phenome-wide scan to discover gene-disease associations. Bioinformatics, 26, 1205–1210.

Gagliano Taliun, S.A. et al. (2020) Exploring and visualizing large-scale genetic associations by using PheWeb. Nat. Genet., 52, 550–552.

Gauderman, W.J. et al. (2007) Testing association between disease and multiple SNPs in a candidate gene. Genet. Epidemiol., 31, 383–395.

Global Lipids Genetics Consortium et al. (2013) Discovery and refinement of loci associated with lipid levels. Nat. Genet., 45, 1274–1283.

Ionita-Laza, I. et al. (2013) Sequence kernel association tests for the combined effect of rare and common variants. Am. J. Hum. Genet., 92, 841–853.

Kamat, M.A. et al. (2019) PhenoScanner V2: an expanded tool for searching human genotype–phenotype associations. Bioinformatics, 35, 4851–4853.

Korotkevich, G. et al. (2019) Fast gene set enrichment analysis. bioRxiv, doi 10.1101/060012.

Kwee, L.C. et al. (2008) A powerful and flexible multilocus association test for quantitative traits. Am. J. Hum. Genet., 82, 386–397.

Lamparter, D. et al. (2016) Fast and Rigorous Computation of Gene and Pathway Scores from SNP-Based Summary Statistics. PLoS Comput. Biol., 12, e1004714.

Lee, S. et al. (2014) Rare-variant association analysis: study designs and statistical tests. Am. J. Hum. Genet., 95, 5–23.

de Leeuw, C.A. et al. (2015) MAGMA: generalized gene-set analysis of GWAS data. PLoS Comput. Biol., 11, e1004219.

Li, M.-X. et al. (2011) GATES: a rapid and powerful gene-based association test using extended Simes procedure. Am. J. Hum. Genet., 88, 283–293.

Liu, J.Z. et al. (2010) A versatile gene-based test for genome-wide association studies. Am. J. Hum. Genet., 87, 139–145.

McCullagh, P. and Nelder, J.A. (1983) Generalized Linear Models Chapman and Hall.

McKusick, V.A. (1998) Mendelian inheritance in man: a catalog of human genes and genetic disorders JHU Press.

McNair, W.P. et al. (2004) SCN5A mutation associated with dilated cardiomyopathy, conduction disorder, and arrhythmia. Circulation, 110, 2163–2167.

Nielsen, J.B. et al. (2018) Biobank-driven genomic discovery yields new insight into atrial fibrillation biology. Nat. Genet., 50, 1234–1239.

Nikpay, M. et al. (2015) A comprehensive 1,000 Genomes-based genome-wide association meta-analysis of coronary artery disease. Nat. Genet., 47, 1121–1130.

Ochoa, D. et al. (2021) Open Targets Platform: supporting systematic drug-target identification and prioritisation. Nucleic Acids Res., 49, D1302–D1310.

Pedregosa, F. et al. (2011) Scikit-learn: Machine Learning in Python. J. Mach. Learn. Res., 12, 2825–2830.

Purcell, S. et al. (2007) PLINK: A Tool Set for Whole-Genome Association and Population-Based Linkage Analyses. Am. J. Hum. Genet., 81, 559–575.

Qin, D. et al. (2019) Atrial Fibrillation-Mediated Cardiomyopathy. Circ. Arrhythm. Electrophysiol., 12, e007809.

Rahman, F. et al. (2014) Global epidemiology of atrial fibrillation. Nat. Rev. Cardiol., 11, 639–654.

Raudvere, U. et al. (2019) g:Profiler: a web server for functional enrichment analysis and conversions of gene lists (2019 update). Nucleic Acids Res., 47, W191–W198.

Roselli, C. et al. (2018) Multi-ethnic genome-wide association study for atrial fibrillation. Nat. Genet., 50, 1225–1233.

Selcen, D. and Engel, A.G. (2004) Mutations in myotilin cause myofibrillar myopathy. Neurology, 62, 1363–1371.

Storey, J.D. and Tibshirani, R. (2003) Statistical significance for genomewide studies. Proc. Natl. Acad. Sci. U. S. A., 100, 9440–9445.

Subramanian, A. et al. (2005) Gene set enrichment analysis: a knowledge-based approach for interpreting genome-wide expression profiles. Proc. Natl. Acad. Sci. U. S. A., 102, 15545–15550.

Svishcheva, G.R. et al. (2019) Gene-based association tests using GWAS summary statistics. Bioinformatics, 35, 3701–3708.

Tucker, N.R. et al. (2020) Transcriptional and Cellular Diversity of the Human Heart. Circulation, 142, 466–482.

Wang, M. et al. (2017) COMBAT: A Combined Association Test for Genes Using Summary Statistics. Genetics, 207, 883–891.

Welter, D. et al. (2014) The NHGRI GWAS Catalog, a curated resource of SNP-trait associations. Nucleic Acids Res., 42, D1001–6.

Wu, M.C. et al. (2011) Rare-variant association testing for sequencing data with the sequence kernel association test. Am. J. Hum. Genet., 89, 82–93.

Yang, J. et al. (2010) Common SNPs explain a large proportion of the heritability for human height. Nat. Genet., 42, 565–569.

Zhan, X. et al. (2016) RVTESTS: an efficient and comprehensive tool for rare variant association analysis using sequence data. Bioinformatics, 32, 1423–1426.

