## Supplementary Tables and Figures for "ExPheWas: a browser for gene-based pheWAS associations"

### Supplementary Material

Marc-André Legault, Louis-Philippe Lemieux Perreault, Marie-Pierre Dubé

### Supplementary Figures

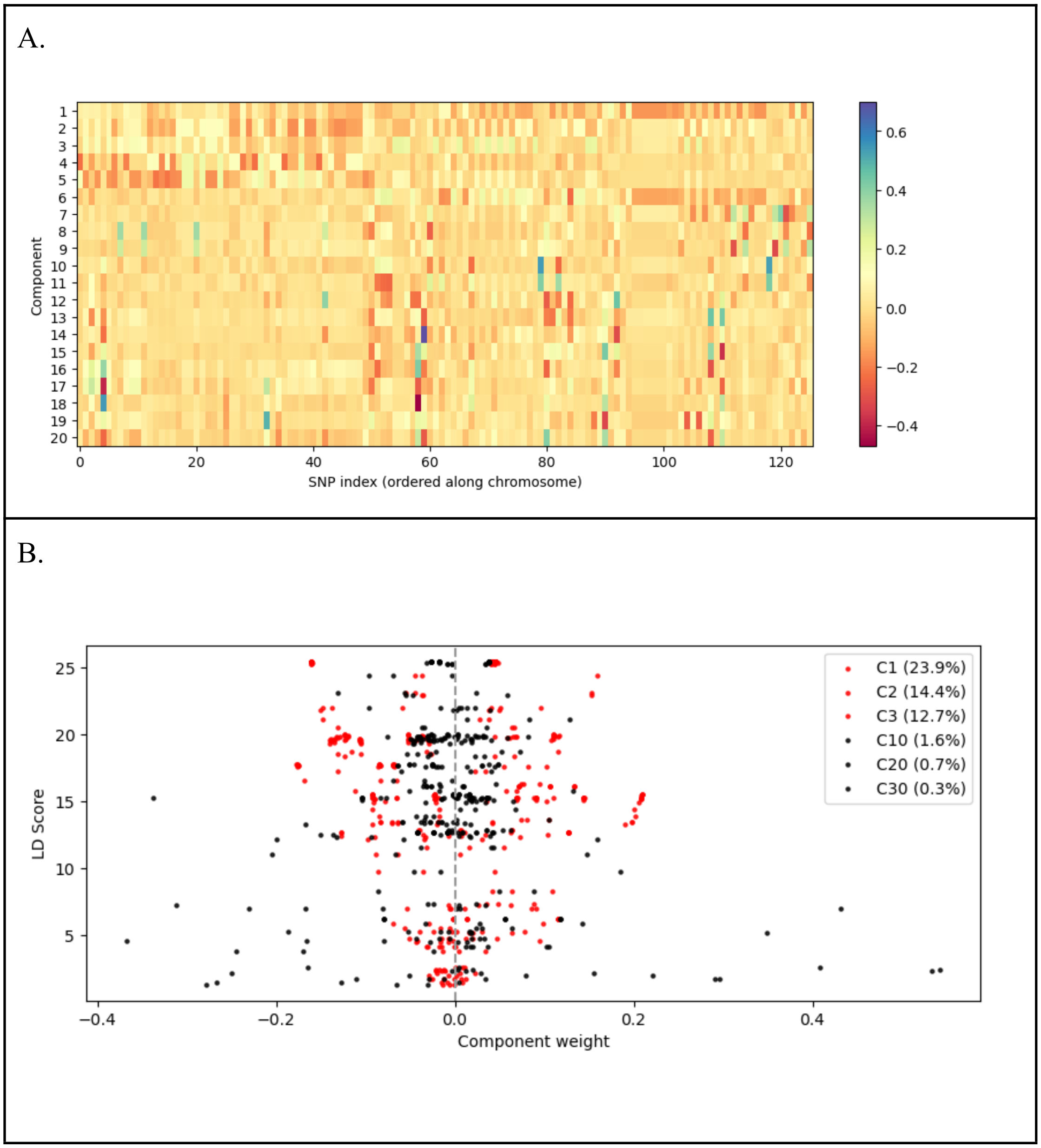

***Supplementary Figure 1. Visualisation of the PCA components along with their position on the chromosome and LD score.*** ***A.*** *PCA weights for the first 20 principal components computed in 503 unrelated European participants of the 1000 Genomes Project (phase 3) using genotypes for 126 common bi-allelic variants at the PCSK9 gene locus.* ***B.*** *LD Score for the variants with respect to component weights. We selected representative components explaining a large portion of the variance in genotypes (components 1 to 3, in red) and components with smaller eigenvalues explaining a smaller proportion of the variance (components 10, 20 and 30, in black). We notice that the first components attribute larger weights to variants with higher LD scores whereas the later components attribute higher weights to variants with smaller LD scores.*

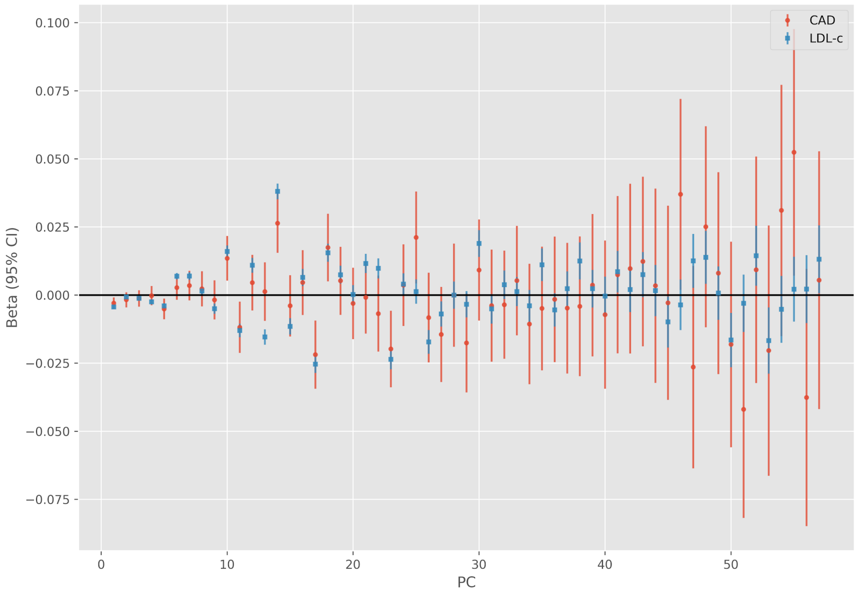


***Supplementary Figure 2****. Marginal association between genetic PCs based on genotypes at the PCSK9 locus and low density lipoprotein cholesterol (blue) and coronary artery disease (red).*

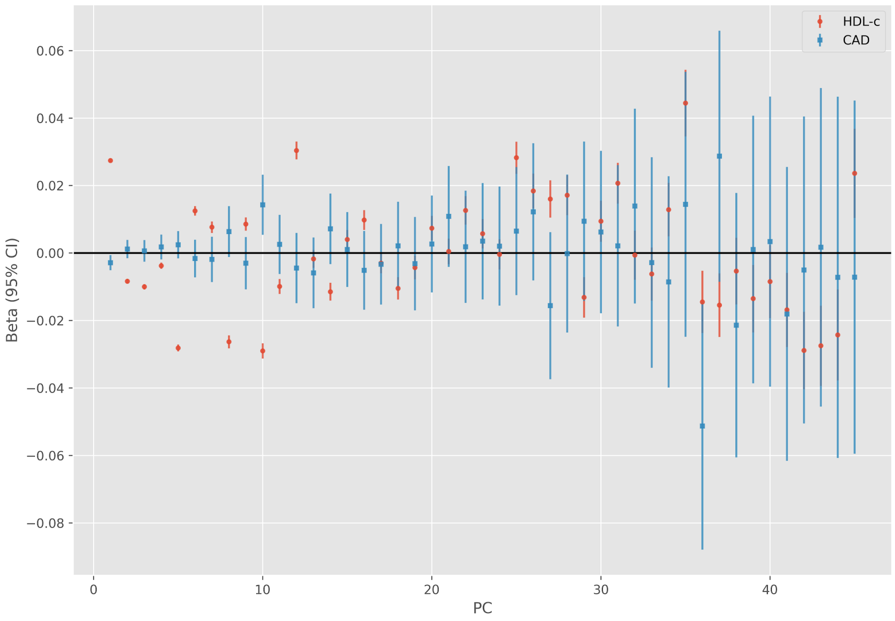


***Supplementary Figure 3***. *Marginal association between genetic PCs based on genotypes at the CETP locus and low density lipoprotein cholesterol (blue) and coronary artery disease (red).*

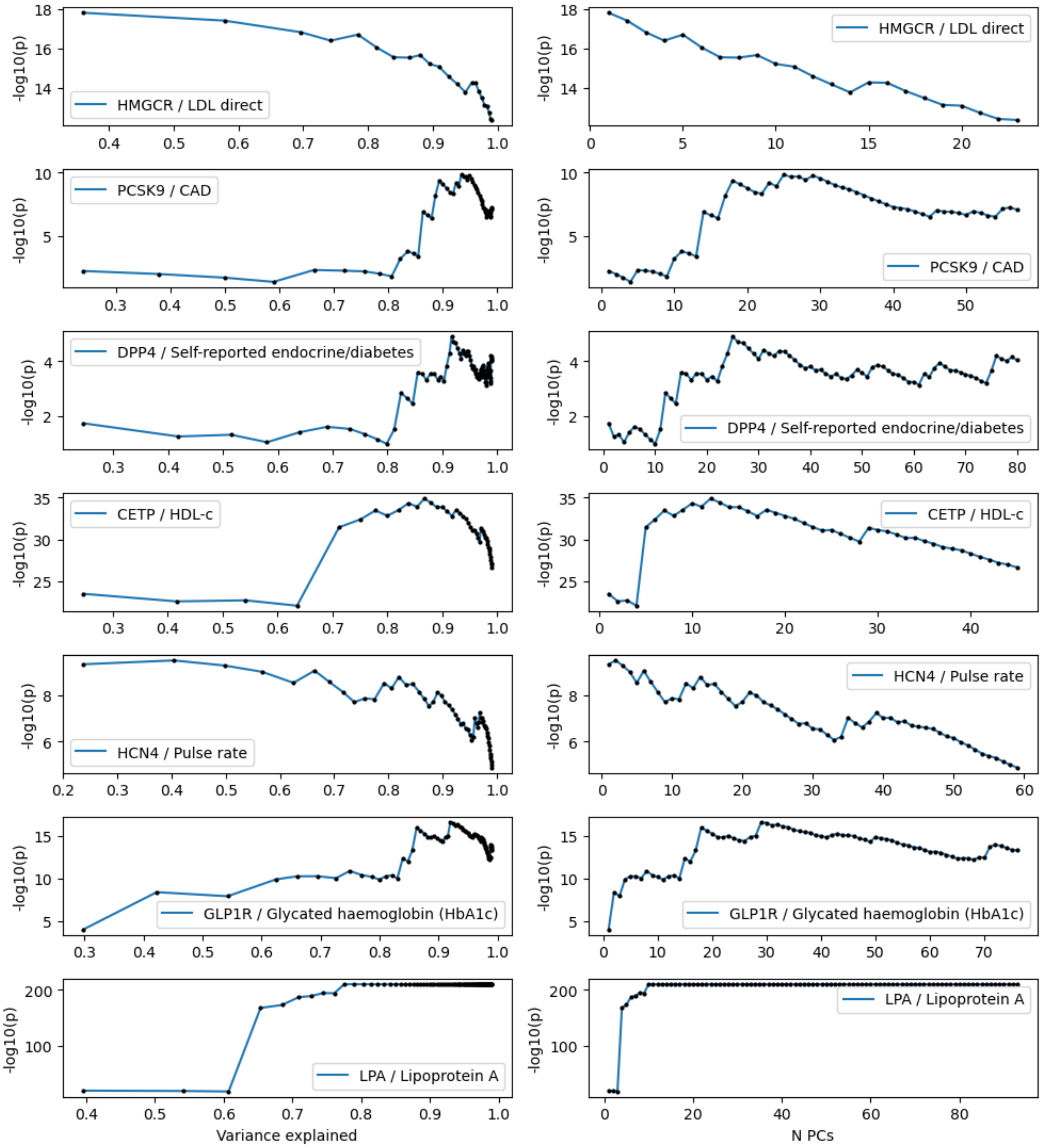


***Supplementary Figure 4**. Association p-value for selected drug target genes and phenotypes for different choices of included PCs. The choice of included PCs is expressed as a proportion of cumulative variance explained (left) and as the actual number of included PCs (right). To*

*avoid saturation of the association p-values at 0, some phenotypes were subsampled by randomly selecting individuals.*

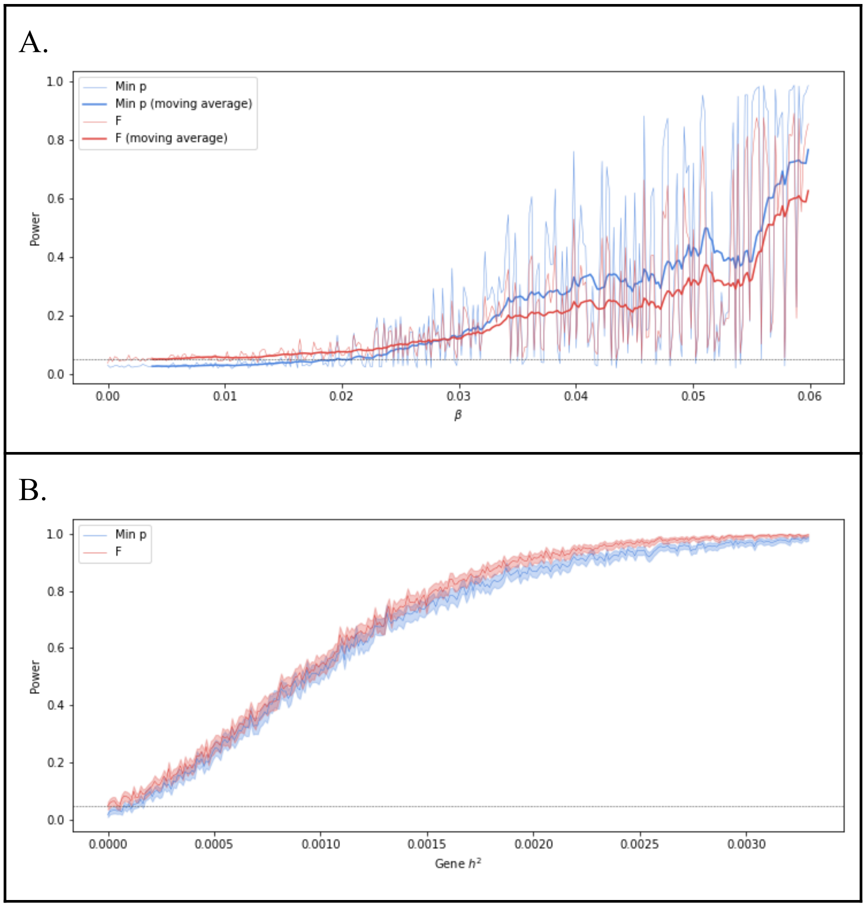


***Supplementary Figure 5. Results from 1000 simulation replicates of 20,000 randomly sampled individuals assessing the power of the PC-based association model and the minimum linear regression p-value within gene boundaries approach. A.*** *We used a simulation model with a single randomly selected causal variant of increasing effect size (β). The wider lines represent a 20 points moving average.* ***B.*** *We used a random effects model where every variant at the locus has an effect drawn from a normal distribution collectively explaining the simulated heritability ($h^2$). The shaded region represents the 95% confidence interval for the power estimate. When taking the minimum p-value in the region we rejected the null hypothesis using a bonferroni adjusted p-value threshold based on the number of tested*

*variants and a α = 0.05 level. Both simulation models include the null model (β or $h^2$ = 0) and the dashed line represents the nominal type I error rate of 0.05.*

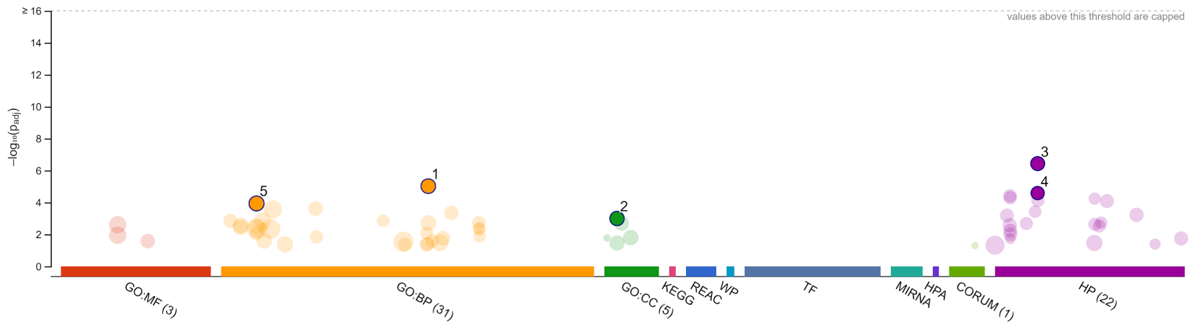


| ID | Source | Term ID | Term Name | $p_{adj}$ (query_1) |
|---|---|---|---|---|
| 1 | GO:BP | GO:0060048 | cardiac muscle contraction | $9.515 \times 10^{-6}$ |
| 2 | GO:CC | GO:0030018 | Z disc | $1.007 \times 10^{-3}$ |
| 3 | HP | HP:0005115 | Supraventricular arrhythmia | $3.684 \times 10^{-7}$ |
| 4 | HP | HP:0005110 | Atrial fibrillation | $2.580 \times 10^{-5}$ |
| 5 | GO:BP | GO:0006941 | striated muscle contraction | $1.151 \times 10^{-4}$ |

**version** e102_eg49_p15_7a9b4d6
**date** 2/10/2021, 11:29:21 AM
**organism** hsapiens

g:Profiler

***Supplementary Figure 6****. Results from a gene set enrichment analysis using g:Profiler for the 137 atrial fibrillation-associated genes with $q \leq 0.01$. Some representative top enrichments are identified on the plot. The full results are available from the g:Profiler website at [https://biit.cs.ut.ee/gplink/l/hMthhT44Rl](https://biit.cs.ut.ee/gplink/l/hMthhT44Rl)*

*A.*

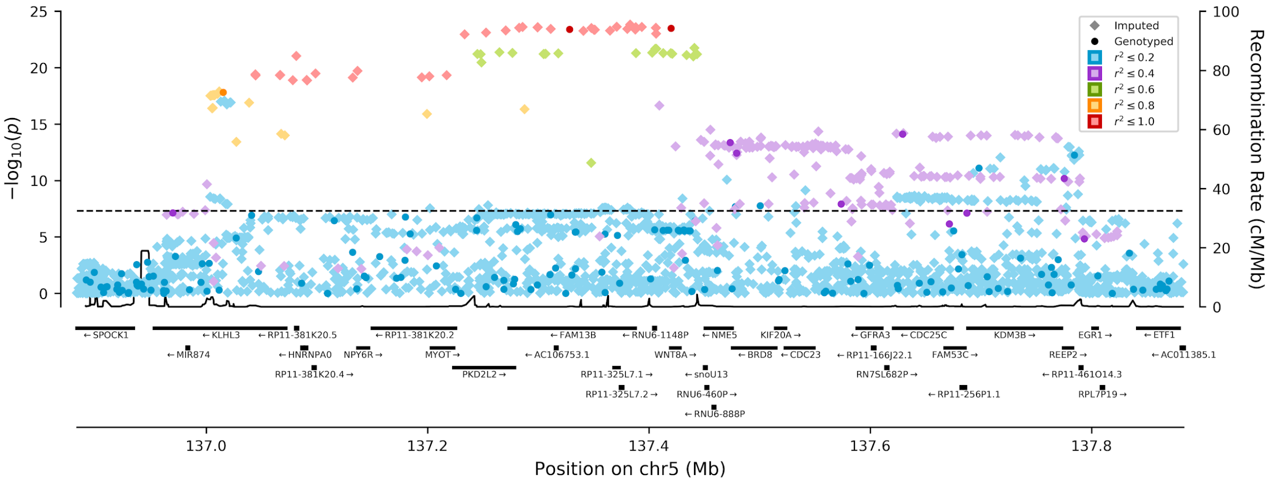
−log10(p)
Recombination Rate (cM/Mb)
Position on chr5 (Mb)
Imputed
Genotyped
r² ≤ 0.2
r² ≤ 0.4
r² ≤ 0.6
r² ≤ 0.8
r² ≤ 1.0
←SPOCK1
←KLHL3
←RP11-381K20.5
←RP11-381K20.2
←FAM13B
←RNU6-1148P
←NME5
KIF20A→
←GFRA3
←CDC25C
KDM3B→
EGR1→
←ETF1
←MIR874
←HNRNPA0
NPY6R→
MYOT→
←AC106753.1
WNT8A→
←BRD8
←CDC23
←RP11-166J22.1
FAM53C→
REEP2→
←AC011385.1
RP11-381K20.4→
PKD2L2→
RP11-325L7.1→
←snoU13
RN7SL682P→
←RP11-461O14.3
RP11-325L7.2→
RNU6-460P→
←RP11-256P1.1
RPL7P19→
←RNU6-888P

*B.*

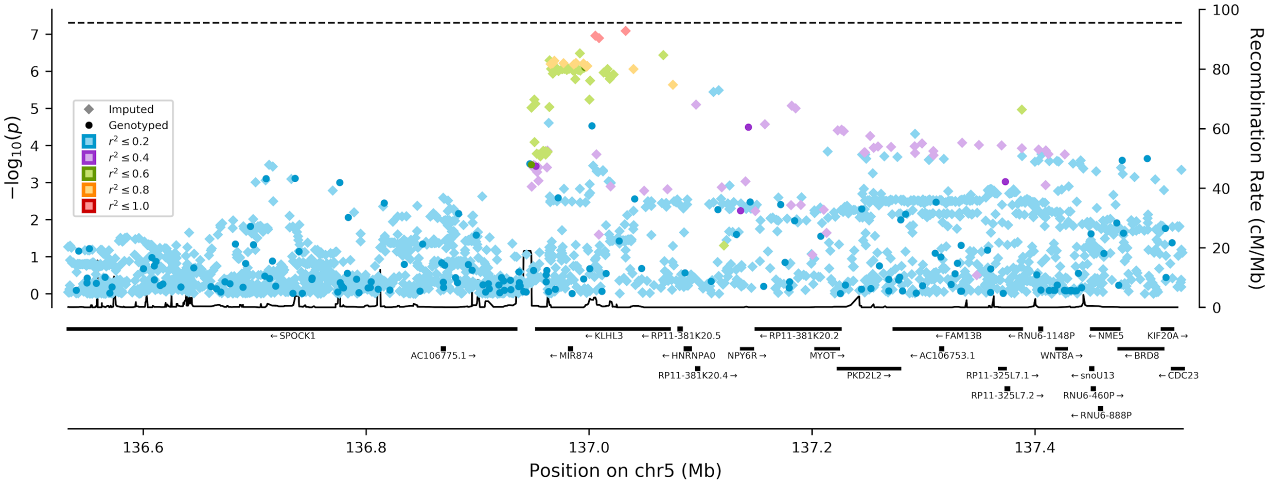
−log10(p)
Recombination Rate (cM/Mb)
Position on chr5 (Mb)
Imputed
Genotyped
r² ≤ 0.2
r² ≤ 0.4
r² ≤ 0.6
r² ≤ 0.8
r² ≤ 1.0
←SPOCK1
AC106775.1→
←KLHL3
←MIR874
←RP11-381K20.5
←HNRNPA0
RP11-381K20.4→
NPY6R→
←RP11-381K20.2
MYOT→
PKD2L2→
←FAM13B
←AC106753.1
RP11-325L7.1→
RP11-325L7.2→
←RNU6-1148P
WNT8A→
←NME5
←snoU13
RNU6-460P→
←RNU6-888P
KIF20A→
←BRD8
←CDC23

*C.*

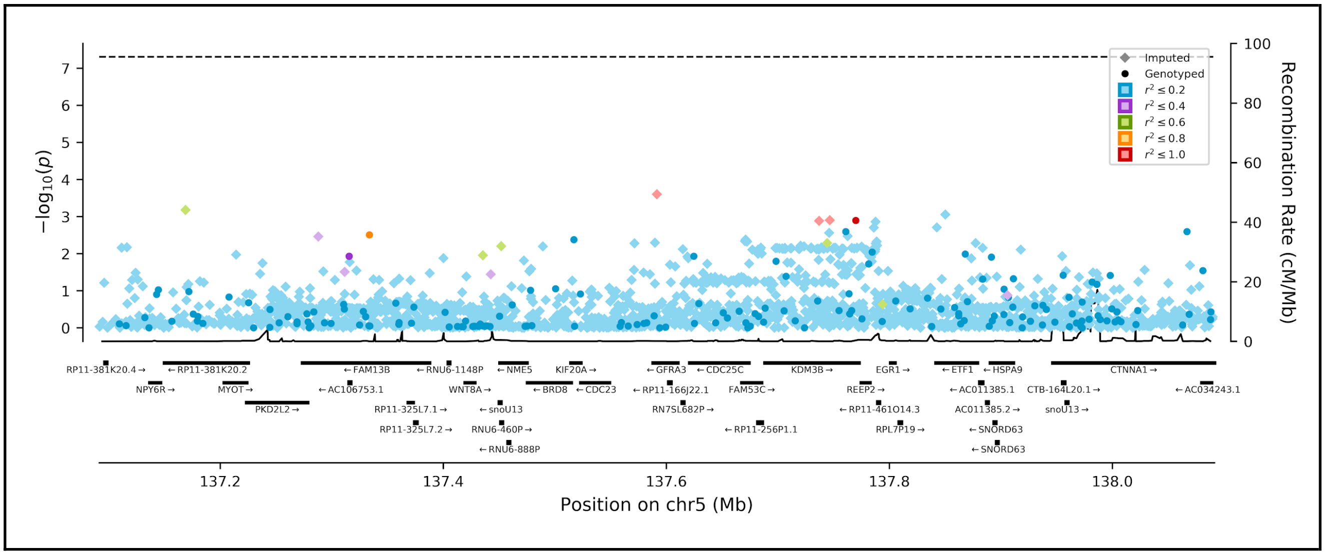


***Supplementary Figure*** *7*. ***Stepwise forward conditional analysis of atrial fibrillation associated variant at the chr5:136,883,078-137,883,078 (GRCh37) locus in the UK Biobank.*** ***A.*** *Initial scan conditional only on age, sex and the first 10 PCs.* ***B.*** *Association scan conditional on the lead variant from stage 1, rs148378888.* ***C.*** *Association scan conditional on the lead variants from previous stages: rs148378888 and rs12653760.*

### Supplementary Tables

***Supplementary Table 1***. *Summary of the included continuous variables and transformations used to obtain approximately normally distributed variables.*

*[See Excel file]*

***Supplementary Table 2***. *Definition of the algorithmically-defined outcomes.*

| Outcome | Definition* |
|---|---|
| **Death outcomes** | |
| Any death | Any entry in the death records variable #40001 |
| Cardiovascular death | Any “I” code as the primary cause of death |
| Coronary artery disease death | I20-I25 as the primary cause of death |
| **Cardiovascular composite outcomes** | |
| Myocardial infarction | ICD9 codes: 410, 412, 411.0, 429.79<br>ICD10 codes: I21, I22, I23, I25.2<br>in the hospitalization or death records |
| Percutaneous coronary intervention / Coronary artery bypass graft (PCI/CABG) | OPCS procedure codes: K40, K41, K42, K43, K44, K45, K46, K49, K50, K75 |
| Unstable angina | I20.0 code as the primary reason for hospitalization or cause of death |
| Any angina | ICD9 code 413 or ICD10 code I20 in the hospitalization or death records |
| Coronary artery disease | ICD9 codes: 410-414 except for aneurysms (414.1)<br>ICD10 codes: I20-I25<br>in the hospitalization or death records or<br>operation codes for PCI/CABG as previously defined |
| Heart failure | ICD9 codes: 425, 428<br>ICD10 codes: I42, I50<br>in the hospitalization or death records |

** Unless otherwise specified, codes were taken in both the primary and secondary reasons for hospitalization, but only the primary cause of death was used.*

***Supplementary Table 3***. *Selected drug target genes and phenotypes and the optimal choice in the number of PCs to include to maximise the association strength.*

| Gene (symbol) | Tested phenotype | Associated drug class | Gene length (quantile) | Optimal number of PCs (cumulative variance explained) | *n* PCs explaining 95% of the variance |
|---|---|---|---|---|---|
| 3-hydroxy-3-methylglutaryl-CoA reductase (*HMGCR*) | LDL cholesterol | Statins (*e.g.* atorvastatin, simvastatin) | 26 kb (0.50) | 1 (36%) | 14 |
| Proprotein convertase subtilisin/kexin type 9 (*PCSK9*) | Coronary artery disease | PCSK9 inhibitors (*e.g.* alirocumab, evolocumab) | 25 kb (0.50) | 25 (93%) | 30 |
| Dipeptidyl-peptidase 4 (*DPP4*) | Self-reported endocrine/diabetes | DPP4 inhibitors (*e.g.* linagliptin) | 82 kb (0.79) | 25 (92%) | 37 |
| Cholesteryl ester transfer protein (*CETP*) | High density lipoprotein cholesterol | CETP inhibitors (*e.g.* anacetrapib, evacetrapib, dalcetrapib) | 22 kb (0.46) | 12 (88%) | 23 |
| Hyperpolarization activated cyclic nucleotide-gated potassium channel 4 (*HCN4*) | Pulse rate | Ivabradine | 49 kb (0.67) | 6 (66%) | 33 |
| Glucagon-like peptide 1 receptor (*GLP1R*) | Glycated haemoglobin (HbA1c) | GLP1R agonists (*e.g.* liraglutide, albiglutide) | 39 kb (0.61) | 29 (92%) | 39 |
| lipoprotein, Lp(a) (*LPA*) | Lipoprotein(a) | Reduced by PCSK9 inhibitors, antisense-based therapies in development | 135 kb (0.88) | 25 (90%) | 45 |

***Supplementary Table 4****. Full g:Profiler ontological enrichment results for the genes associated with atrial fibrillation.*

*[See Excel file]*
